# A new role for Biofoundries in rapid prototyping, development, and validation of automated clinical diagnostic tests for SARS-CoV-2

**DOI:** 10.1101/2020.05.02.20088344

**Authors:** Michael A. Crone, Miles Priestman, Marta Ciechonska, Kirsten Jensen, David J. Sharp, Paul Randell, Marko Storch, Paul Freemont

**Affiliations:** London Biofoundry, Imperial College Translation & Innovation Hub, White City Campus, 80 Wood Lane, London, W12 0BZ, UK; Section of Structural and Synthetic Biology, Department of Infectious Disease, Imperial College London, London, SW7 2AZ, UK; UK DRI Care Research and Technology Centre, Imperial College London, Hammersmith Campus, Du Cane Road, London, W12 0NN London, UK; North West London Pathology, Department of Infection and Immunity; Imperial College Healthcare NHS Trust, Charing Cross Hospital Fulham Palace Rd, Hammersmith, London W6 8RF

## Abstract

The SARS-CoV-2 pandemic has shown how the rapid rise in demand for patient and community sample testing, required for tracing and containing a highly infectious disease, has quickly overwhelmed testing capability globally. With most diagnostic infrastructure dependent on specialised instruments, their exclusive reagent supplies quickly become bottlenecks in times of peak demand, creating an urgent need for novel approaches to boost testing capacity. We address this challenge by refocusing the full synthetic biology stack available at the London Biofoundry onto the development of alternative patient sample testing pipelines. We present a reagent-agnostic automated SARS-CoV-2 testing platform that can be quickly deployed and scaled, and that accepts a diverse range of reagents. Using an in-house-generated, open-source, MS2-virus-like-particle-SARS-CoV-2 standard, we validate RNA extraction and RT-qPCR workflows as well as two novel detection assays based on CRISPR-Cas and Loop-mediated isothermal Amplification (LAMP) approaches. In collaboration with an NHS diagnostic testing lab, we report the performance of the overall workflow and benchmark SARS-CoV-2 detection in patient samples via RT-qPCR, CRISPR-Cas, and LAMP against clinical test sets. The validated RNA extraction and RT-qPCR platform has been installed in NHS diagnostic labs with a testing capacity of 1000 samples per day and now contributes to increased patient sample processing in the UK while we continue to refine and develop novel high-throughput diagnostic methods. Finally, our workflows and protocols can be quickly implemented and adapted by members of the Global Biofoundry Alliance and the wider scientific and medical diagnostics community.

## Introduction

Following the report of a case in Wuhan on 31^st^ December 2019, the rapid spread and highly infectious nature of the newly emerged coronavirus has resulted in a worldwide pandemic, as declared by the World Health Organisation (WHO) on 11^th^ March 2020 [1]. The causative agent of COVID-19 has been classified as severe acute respiratory syndrome coronavirus 2 (SARS-CoV-2) and is closely related to the severe acute respiratory syndrome (SARS) and Middle East Respiratory Syndrome (MERS) coronaviruses which were responsible for outbreaks in 2003 and 2012, respectively [2]. As of April 29, there have been 3 100 000 SARS-CoV-2-confirmed cases worldwide, with 215 000 deaths in 185 countries [3]. The fast rate of SARS-CoV-2 human-to-human transmission has resulted in an unprecedented need for diagnostic testing, placing a great strain on public health departments in every country. Diagnostic testing is essential not only for the identification of infection in patients but also for tracking and containment of viral spread within communities and worldwide, testing of unresolved cases, and daily screening of medical frontline workers. It has become apparent that existing diagnostic infrastructure and associated reagent supply lines cannot be scaled to support sufficient testing in the early weeks and months of this pandemic in any country.

Automated workflows are highly preferable over manual protocols to achieve meaningful throughput, diagnostic precision, and to exclude human error from the sample processing pipeline. Typical automated systems such as the Roche cobas^®^ unit can process hundreds of samples per day with minimal staff support, while ensuring uniform processing and sample tracking. As with other similar automated diagnostic testing platforms, they are costly and not available in the numbers needed to process hundreds of thousands of daily samples in the UK. Furthermore, they currently suffer from severe reagent supply shortages reducing their theoretical platform capacity to an insufficient practical testing capacity in the UK as in most countries. Thus, an urgent need has arisen for the adaptation of alternative automated liquid handling platforms and diagnostic test approaches and workflows, ideally designed in an open and modular way to allow for diversifying reagent supply away from mainstream and overstretched reagent sources.

Many research institutions around the world have established non-commercial Biofoundries, that offer an integrated infrastructure including state-of-the-art automated high-throughput (HT) equipment to enable the design-build-test cycle for large scale experimental designs in synthetic biology [4]. The unique combination of HT infrastructure with technical expertise in molecular biology, analytics, automation, engineering, and software development provides an excellent, self-sufficient, and agile capability to quickly establish platforms for prototyping biological testing standards and developing liquid-handling workflows, such as those needed for automated diagnostic testing of SARS-CoV-2. Once new workflows are validated, the Global Biofoundry Alliance provides an established network and collaborative culture which allows their quick implementation across the world. In the London Biofoundry, we have rapidly re-configured our existing liquid handling infrastructure to establish a HT SARS-CoV-2 testing workflow that can be implemented immediately into existing frontline diagnostic labs. We have also established two novel workflows using CRISPR-Cas and LAMP which would allow increased future testing capacity.

To generate a standard for our workflow development, benchmarking, and validation, we also engineered non-infectious synthetic virus-like particles (VLP). ‘Armoured RNA’ is a non-infectious RNA virus surrogate consisting of an MS2 bacteriophage capsid containing an RNA template of choice [5]. It has been employed as a diagnostic reference tool for the detection of respiratory viruses such as Influenza A and B as well as SARS-CoV [6], [7]. The particles are non-infectious and do not require specialist laboratories or equipment as is the case for live patient samples. Furthermore, they are nuclease resistant, have been shown to be highly stable in plasma, nasopharyngeal secretions, faeces, and water, and simulate the presence of a real viral target [7]. This makes them an excellent choice for the development and validation of diagnostic workflows, including RNA extraction and detection of transcripts by RT-qPCR, CRISPR-Cas systems, or colorimetric LAMP [8].

Here, we produced and characterised a VLP standard simulating SARS-CoV-2 by containing the genomic RNA segments complementary to the N protein N1, N2, and N3 primer-probe sets specified by the CDC 2019-Novel Coronavirus (2019-nCoV) Real-Time RT-PCR Diagnostic Panel [9][10]. We show that these provide good sensitivity and thus a low limit of detection in RT-qPCR assays. We used the SARS-CoV-2-VLPs as a semi-quantitative copy number standard and as a processing control in the absence of patient samples to design and optimise an automated nucleic acid test (NAT) diagnostic workflow encompassing viral RNA extraction and one-step RT-qPCR. The optimised workflow has been validated using patient samples and results shows high correlation with accredited diagnostic laboratory test results. Finally, we have modularised the workflow to anticipate deficits in the reagent supply chain or qPCR equipment. We have implemented multiple off-the-shelf RNA extraction kits, assessed the quality of several RT-qPCR reagent suppliers, and employed CRISPR-Cas diagnostic and colorimetric isothermal amplification systems, as alternative SARS-CoV-2 detection methods.

As of the date of submission, the UK is processing approximately 85 000 samples per day [11]. The diagnostic pipeline outlined here provides a modular approach to maximising testing capability using an alternative automation platform with several options for RNA extraction, RT-qPCR master mixes, and qPCR-free workflows. Furthermore, by automating the workflow, we report an average sample processing rate of approximately 1 000 samples per platform per day which can be modified and scaled to 4 000 samples per day. Another benefit is that each platform only requires a small laboratory bench and is easily portable for testing in areas of low population density. With our new workflow validated and implemented at two NHS diagnostic laboratories, our work has helped increase the current testing capacity in London and will provide support and validation for Biofoundries and interested laboratories globally.

## Results

### VLP preparation and characterisation

Recombinant MS2 bacteriophage VLPs carrying the SARS-CoV-2 N gene were produced in *E. coli* from an expression plasmid using protocols described previously and modified to transcribe and package the primer-probe target sites for the CoV-2 N protein (Fig 1a)[12]. The assembled MS2-SARS-CoV-2 VLPs were purified and treated with DNase and RNase to ensure the preparation was free from template DNA and RNA contamination. The purity of the sample was analysed by SDS-PAGE (Fig 1b). We determined the VLP size distribution using dynamic light scattering (DLS) to be approximately 27 nm (Fig 1c), which matched well with a previously well characterized MS2 VLP construct [12]. Next, we employed digital droplet PCR (ddPCR) to obtain absolute quantities of three serial dilutions of VLPs. Heat lysis has been shown to be effective in releasing RNA from armoured RNA VLPs at levels comparable to commercially available extraction kits [7]. RNA encoding the CoV-2 N protein was extracted from the VLPs at 95°C for five minutes, followed by amplification using the N1 primer-probe set. The released RNA was serially diluted and absolute quantification was performed. The ddPCR quantification method involves partitioning of the sample into thousands of droplets which individually contain single amplification reactions using the N1 probe. VLP concentration can then be derived using Poisson distribution statistics to determine absolute particle concentration in each dilution. This method allows for highly accurate and precise sample quantification without the need for a standard curve. We analysed the purified MS2-SARS-CoV-2 VLP absolute concentration in serial ten-fold dilutions, which were found to contain 250, 25 and 2.5 copies per ml respectively (Fig 1d).

**Figure 1:**
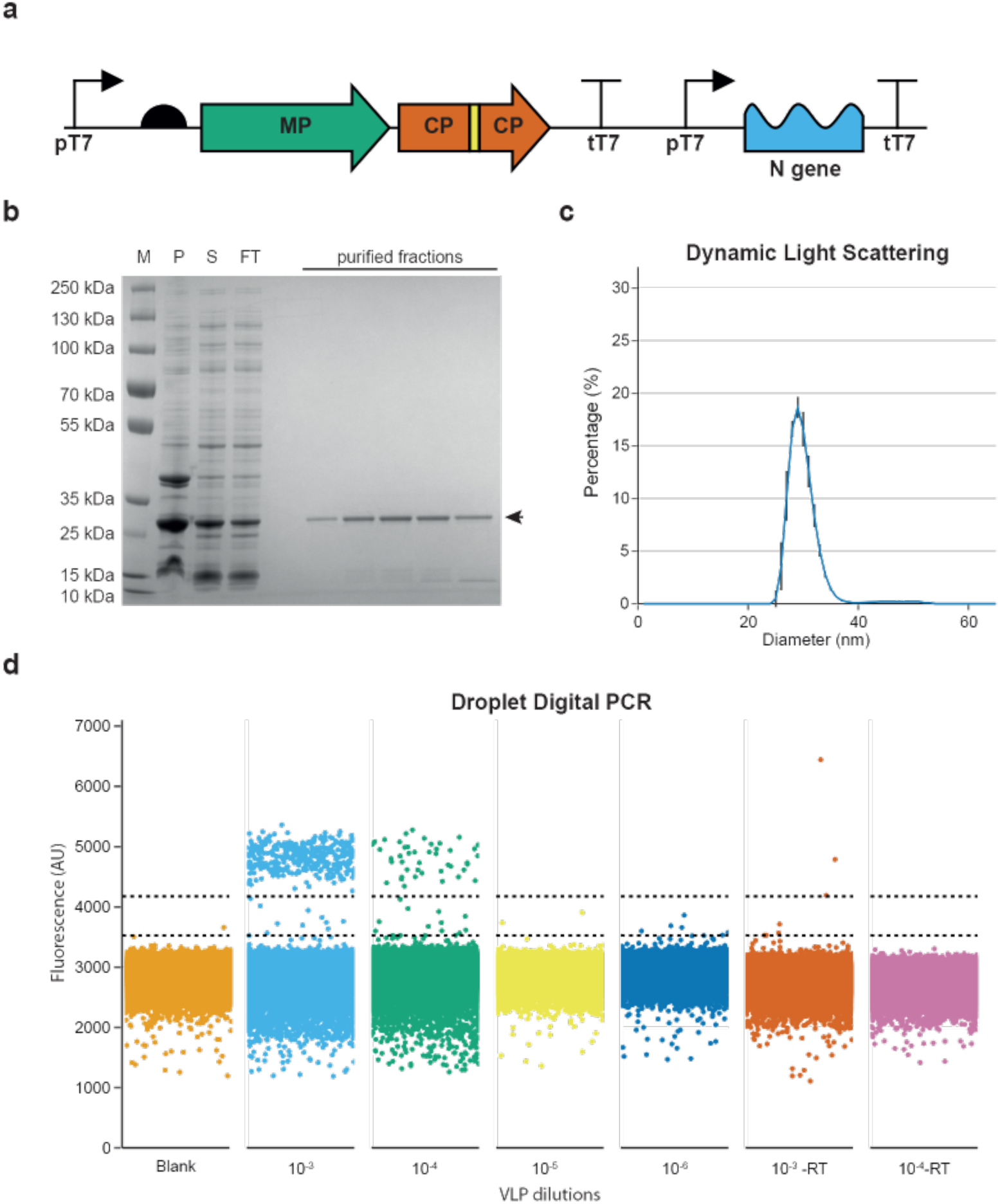
SARS-CoV-2 VLP production and characterisation. (**a**) Schematic of the genetic construct of the engineered MS2 SARS-CoV-2-N gene VLP encompassing the MS2 maturation protein (MP) in green and coat proteins (CP) in orange, linked via His-tag (yellow), under transcriptional control of the T7 promoter and T7 terminator sequences. The SARS CoV-2 N protein RNA is packaged using a downstream *pac site*. (**b**) The VLP constructs were expressed in *E. coli* and purified using HiTrap^®^ TALON^®^ Crude and HiTrap^®^ Heparin columns. SDS-PAGE analysis of the purification steps includes a protein marker (M) followed by pellet (P) and soluble fraction (S) of the cell lysate, followed by the column flow through (FT) and protein elution fractions, where the CP-His-CP dimer (~28 kDa) is indicated by an arrow. (**c**) Purified VLPs were analysed by Dynamic Light Scattering (DLS) which showed a uniform particle population of approximately ~27nm. Error bars represent the standard deviation of three technical replicates. (**d**) Droplet Digital PCR (ddPCR) was performed for absolute quantification of the purified VLPs. Serial dilutions of 1, 10, and 100 thousand-fold of the purified VLPs in the presence and absence of a reverse transcription (RT) step were analysed. Droplets were clustered using a threshold determined using a python implementation of an online tool (http://definetherain.org.uk). Dotted lines represent the cut offs for the positive and negative clusters. Any data points between the two dotted lines are considered droplet “rain”.

### VLP validation as a SARS-CoV-2 standard by RT-qPCR

One-step Quantitative RT-PCR (RT-qPCR) is currently the ‘gold standard’ for detection of nucleic acids in molecular diagnostic tests due to its sensitivity, robustness, dynamic range, HT capability, and affordability. It is the current method of choice for the detection of SARS-CoV-2 in the UK and around the world. In order to demonstrate the utility of MS2-SARS-CoV-2 VLPs as a standard for optimising and validating automated NAT diagnostic workflows, we assessed whether they could be reliably detected via RT-qPCR using the CDC 2019-Novel Coronavirus (2019-nCoV) Diagnostic Panel primer-probe set. To this end, we extracted the SARS-Cov-2 N protein RNA encapsulated by the MS2 VLP using heat lysis from the serial dilutions quantified via ddPCR and performed One-Step RT-qPCR using the TaqPath master mix (Thermo Fisher Scientific). Three biological replicates were completed to assess the robustness of using the MS2-SARS-CoV-2 VLPs as control RNA (Fig 2a). The quantified VLP dilutions were also used to generate a standard curve to aid in assessment of viral RNA purification efficiency and to estimate the limit of detection (LoD) of our automated workflow (Fig 2b).

**Figure 2:**
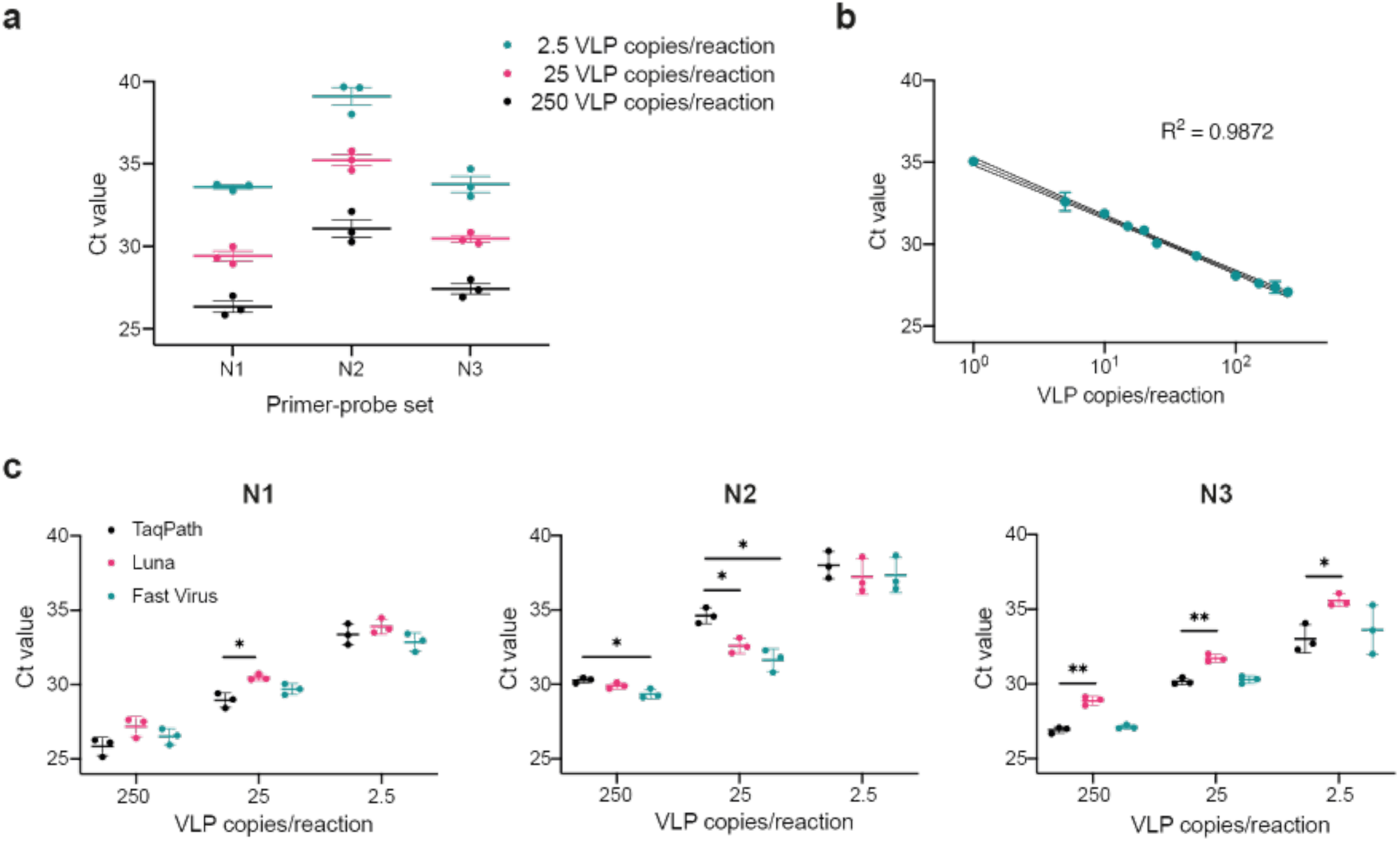
MS2 SARS-CoV-2 N gene VLPs can be detected at low concentrations with multiple target primer-probe sets and qPCR master mixes. (**a**) VLP dilutions of 2.5, 25, and 250 copies per reaction were analysed by One-Step RT-qPCR using the CDC primer-probe sets N1, N2, and N3 with the TaqPath master mix (ThermoFisher) and reported as Ct values. (**b**) A Ct value standard curve for VLP concentrations of 1, 5, 10, 15, 20, 25, 50, 100, 150, 200, and 250 VLP copies per reaction was determined using the N1 primer-probe set and the TaqPath master mix. (**c**) VLP dilutions of 250, 25, and 2.5 copies per reaction were analysed using the TaqPath, Luna Universal (NEB), and Fast Virus (ThermoFisher) RT-qPCR master mixes with the N1, N2, and N3 CDC primer-probe sets. All measurements in (a) and (c) are reported as mean ± standard error (SE) of three biological replicate experiments. Measurements in (b) are reported as mean ± standard deviation (SD) and are representative of two biological replicate experiments. Statistical difference between the TaqPath and Luna as well as TaqPath and Fast Virus master mix Ct values was analysed using an unpaired t-test with (*) indicating *p*<.05 and (**) *p*<.01.

The current unprecedented demand for the one-step RT-qPCR master mix to detect SARS-CoV-2 may result in disruption to the laboratory reagent supply chain. Here we used our MS2-SARS-CoV-2 standards to demonstrate modularity of the one-step RT-qPCR detection method by comparing detection reproducibility between three commercially available master mixes using the CDC Diagnostic Panel N1, N2 and N3 primer-probe sets. We report that Ct values achieved using the Virus Fast One-Step (Thermo Fisher Scientific) master mix closely matched those generated with the gold standard TaqPath master mix from the same supplier (Fig 2 c). Ct values obtained using the Luna Universal RT-qPCR kit supplied by New England Biolabs differed slightly from the TaqPath master mix. While all three primer-probe sets achieved similar Ct results, N1 produced the lowest Ct values with least variability, as previously reported by Vogels et al. [13]. The N2 primer-probe set produced higher Ct values and exhibited more variability between replicates for all three RT-qPCR master mix options. Based on its higher sensitivity, we chose the N1 primer-probe set for validation of our RNA extraction and virus detection workflows.

### CRISPR Cas13 as an alternative to one-step RT-qPCR

To further expand the modular nature of the automated platform, we assayed an alternative to the standard RT-qPCR detection method by employing a novel CRISPR Cas13 NAT. This approach, based on the specific high-sensitivity enzymatic reporter unlocking (SHERLOCK) method, was designed to identify and amplify target sequences of the CoV-2 N gene RNA packaged within the MS2-SARS-CoV-2 VLP [14]. Briefly, similar to RT-qPCR, the initial step of this method relies on the reverse transcription and amplification of the target RNA. Here, we employed the above-mentioned CDC diagnostic primer sets N1, N2, and N3 (with forward primers 5’ extended with a T7 promoter sequence) together with a one-step RT enzyme mix to generate cDNA from N gene RNA released from the VLPs. However, unlike qPCR, the subsequent step includes transcription of the amplified segment to RNA and incubation with a guide RNA (gRNA), in this case complementary to the N region of CoV-2, together with a purified recombinant Cas13a protein derived from *Leptotrichia wadei* (LwCas13a). Upon recognition and binding of the target sequence by gRNA, Cas13 exhibits RNase activity not only for this complex, but also collateral activity for any RNA in its vicinity. Taking advantage of this non-specificity, a quenched fluorescent probe can be added and subsequently cleaved by activated Cas13, thus generating a quantitative signal which can be detected using a standard fluorescence microplate reader. By applying this technology, we assayed MS2-SARS-CoV-2 RNA released through heat lysis and were able to detect the CoV-2 RNA sequence at 2500, 250 and 2.5 VLP copies per reaction (Fig 3). This low limit of detection was comparable to all of the one-step RT-qPCR master mixes reported above, although viral load quantification is difficult. Thus, we propose this method may be used to substitute current RT-qPCR diagnostic workflows, as it does not require qPCR equipment and is highly amenable to HT automated workflows. Furthermore, it enables 10- to 100-fold increased throughput when assembled in high-density assay plates, with accurate diagnostic test readout available in just a few minutes using a standard fluorescence microplate reader. Another advantage of our CRISPR workflow is the possibility of identifying specific viral serotypes in a multiplexed strain-specific diagnostic which would provide additional information for clinical management.

**Figure 3:**
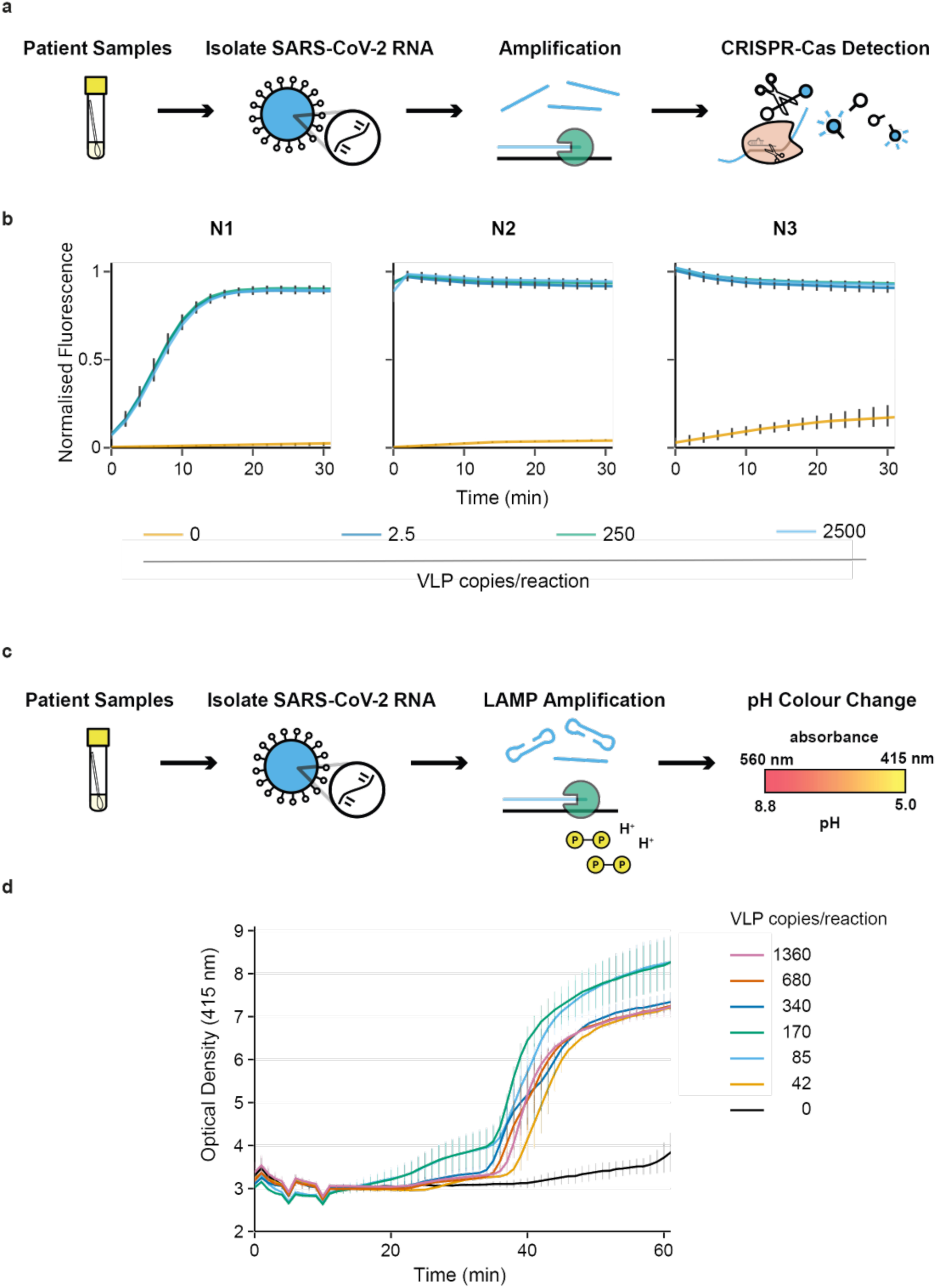
SARS-CoV-2 N-gene target RNA detection by CRISPR-Cas and LAMP diagnostic systems. (**a**) Schematic of the CRISPR-Cas13 nucleic acid detection workflow from patient samples. Viral RNA is amplified using target-specific primers and detected with target-specific gRNA activating Cas13 to collaterally cleave a fluorescent probe. (**b**) CDC N1, N2, and N3 primer sets were employed to amplify the N gene RNA released from MS2-SARS-CoV-2 VLPs at 2.5, 250, and 2500 copies per reaction. The CRISPR-Cas13 detection time-course was analysed using a fluorescence microplate reader. Error bars represent the standard error of the mean of three independent amplification replicates and four CRISPR detection technical replicates. (**c**) Schematic of the Loop-Mediated Isothermal Amplification (LAMP) diagnostic workflow using target-specific LAMP primers. The isothermal amplification of a target results in the acidification of the LAMP master mix and a subsequent pH-associated colour change that is detected using a microplate reader. (**d**) Time-course detection of a LAMP reaction using the MS2-SARS-CoV-2 VLPs at 42, 85, 170, 340, 680, and 1360 copies per reaction, performed at 65 °C using the BMG CLARIOstar Plus microplate reader. Error bars represent the standard error of the mean of three independent amplification replicates.

### Colorimetric LAMP as an alternative to one-step RT-qPCR

As an additional alternative detection methodology, we adapted the previously described colorimetric LAMP assay [8] for use with high-throughput automation. In this approach, a pyrophosphate moiety and hydrogen ion are produced for every nucleotide that is incorporated into the PCR product during each amplification step. The release of hydrogen ions results in a pH change which can be visually determined using dyes such as phenol red [15] and by measuring absorbance (Fig 3c). Here, we employed the Labcyte Echo platform to set up LAMP reactions using various concentrations of VLP N gene RNA template in 384-well microplates. These were incubated at 65 °C in a microplate reader and absorbance at 415 nm was measured over time. We demonstrated that the presence of the target sequence can be detected reliably down to at least 42.5 copies per reaction, although viral load quantification using LAMP is not currently possible. The use of automation coupled with the speed and affordability of the LAMP workflow provides an excellent alternative to qPCR diagnostic NATs in a format that is highly amenable to ultra-high-throughput workflows (Fig 3d).

### Automated workflow development and validation by RNA extraction and RT-qPCR

Automation of clinical laboratory diagnostics has been essential to increase sample processing throughput, minimise run times, standardise sample processing, maximise accuracy and reproducibility, and to reduce human error. The unprecedented need for diagnostic testing imposed by the SARS-CoV-2 pandemic has resulted in a bottleneck in sample processing throughput. To increase patient sample turnaround time, we have developed an automated diagnostic workflow including RNA extraction and RT-qPCR using elements of the full synthetic biology technology stack available at the London Biofoundry (Fig 4a). We employed the MS2-SARS-CoV-2 VLPs as a process control to optimise and validate our automated clinical diagnostic workflow encompassing RNA extraction and the RT-qPCR and CRISPR Cas13 detection methods. In order to design full-factorial experiments, track randomised samples, and document optimisation and validation experiments, we used the Riffyn platform in combination with JMP software. VLP RNA extraction was optimized for the Analytik Jena CyBio FeliX liquid handling platform for the standard 96-well plate format using the innuPREP Virus DNA/RNA Kit - FX. We prepared two VLP dilutions of 1000 copies/ml and 10 000 copies/ml in order to simulate viral load amounts found in patient samples [16]. Using the optimised FeliX extraction protocol, we were able to isolate RNA from the test VLP dilutions within 60 minutes. The automated workflow takes advantage of magnetic beads-based nucleic acid extraction and eliminates laborious and time-consuming column-based binding and spinning steps. While we tested three biological replicates of the two dilutions, this workflow is designed for the concurrent processing of 96 samples. We project that employing the FeliX RNA extraction protocol in a 96-well format and using one liquid handling device can result in the processing of 1000 samples in 12hrs, including extra time for reagent and extraction kit refilling and patient sample plate loading. This workflow requires minimal user intervention and is therefore highly scalable.

**Figure 4:**
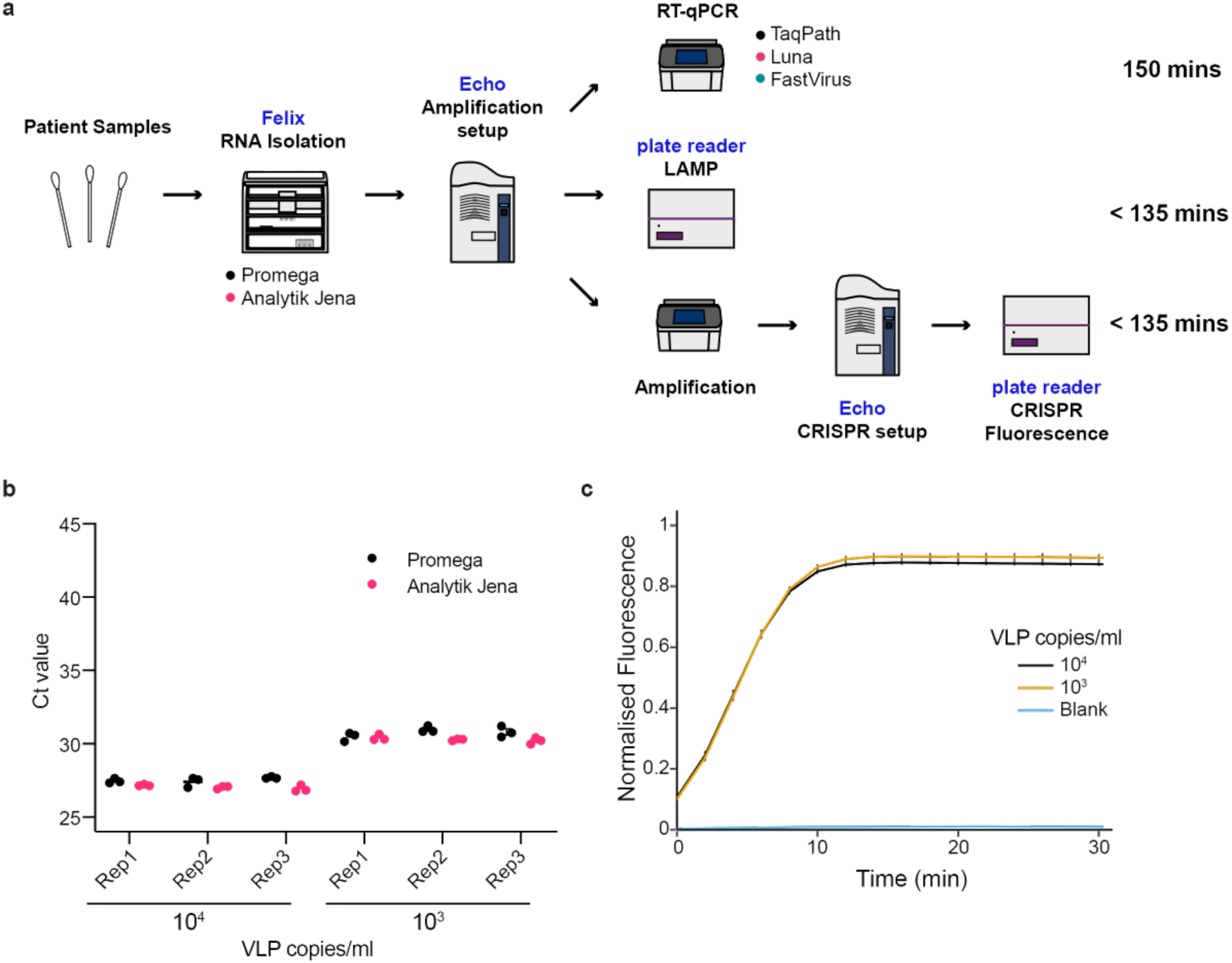
Flexible, high-throughput SARS-CoV-2 testing platform developed using the MS2-SARS-CoV-2 VLP standard. **(a)** Schematic of the modular platform for the detection of SARS-CoV-2. Viral RNA is isolated using the Analytik Jena FeliX liquid handler with the Analytik Jena or Promega RNA extraction kits. Sample RNA is transferred, and detection reactions are set up using the Labcyte Echo platform. These include qPCR, validated for the TaqPath (ThermoFisher), Luna (NEB), and Fast Virus (ThermoFisher) RT-qPCR master mix options, LAMP nucleic acid detection and the CRISPR-Cas13 diagnostic workflow. (**b**) Automated RNA extraction was developed using VLP dilutions of 10^3^ and 10^4^ copies per ml for the Analytik Jena and Promega RNA extraction kits. Efficiency of the extractions using both kits was analysed by RT-qPCR with the CDC N1 primer-probe set using the TaqPath master mix. (**c**) Dilutions of VLPs used for RT-qPCR in (**b**) were analysed using the CRISPR NAT to demonstrate the use of this workflow as an alternative diagnostic option. Error bars in (b) and (c) represent mean ± SE of three biological replicate experiments.

Next, we employed RT-qPCR to determine RNA extraction efficiency. To this end, we used a combination of Riffyn and JMP software to generate and track randomised pick lists for the Labcyte Echo 525 acoustic liquid handling platform, which was used to automate the TaqPath master mix, primer-probe, and sample transfer into 96-well qPCR plates. Plate handling time from RNA extraction to qPCR launch was approximately 10 minutes when using the ECHO 525 followed by a qPCR running time of approximately 70 minutes. The extracted VLP RNA assayed by RT-qPCR resulted in Ct values that approximated those achieved with VLP heat lysis described above for the same concentrations, suggesting a high efficiency of extraction using the automated platform (Fig 4b). Total running time from beginning of the RNA extraction to obtaining RT-qPCR results was approximately 2 hours and 45 minutes.

A key advantage of the Analytik Jena CyBio FeliX liquid handling platform over other commercial dedicated platforms is its programmability. This allows a reagent-agnostic approach to be developed, permitting resilient, robust, and redundant supply chains to be established. To this end, we also implemented and optimised RNA extraction using an alternative kit – Maxwell HT Viral TNA (Promega) – using the same platform. This required minor adjustments to match manufacturer recommendations for optimal volumes and empirically determined mixing steps. Crucially, the hardware is identical, the plasticware is identical, and the output is identical thus requiring no changes to working practice. The same VLP dilutions were used to allow comparison of RNA extraction efficiency between the innuPREP Virus DNA/RNA Kit and the Maxwell HT Viral TNA kit by RT-qPCR (Fig 4b). The Ct values achieved from RNA extracted using the Promega kit are broadly 0.5 cycles higher than those for the Analytik Jena kit (27.52 ± 0.05 and 27.04 ± 0.04 respectively for the high VLP concentration, *p*=2.90 × 10^−7^; and 30.76 ± 0.08 and 30.30 ± 0.05 for the lower concentration, *p*=2.02 × 10^−4^; ± standard error of the mean, paired *t*-test).

In parallel, a CRISPR-Cas workflow was tested for situations where the number of qPCR machines, but not PCR machines, may be a limiting factor. Samples were pre-amplified with one-step RT-PCR master mix and 250 nL of each PCR product was added to 4750 nL of Cas13 gRNA master mix using the Labcyte Echo 550. Reactions reached saturation in positive samples within approximately 10 minutes (Fig 4c). Although this approach slightly lengthens the approximate running time from RNA extraction to obtaining a diagnostic test result to approximately 3 hours, it provides an alternative detection methodology that is more easily scaled than qPCR workflows.

### Validation of the automated platform with patient samples

After demonstrating that the workflow could detect VLPs loaded with SARS-CoV-2 RNA at clinically relevant concentrations, we validated the platform on 173 patient samples obtained from North West London Pathology (NWLP). We compared our extraction (Analytik Jena innuPREP Virus DNA/RNA Kit) and qPCR workflow to that of NWLP at the time (a multiplexed-tandem PCR workflow). We assayed 5 μL of purified RNA from each patient sample (Fig 5a) and showed good correlation (R^2^ = 0.8310) between our results and the test used by NWLP (Fig 5b). Of 173 samples tested, we were able to match 49 positive and 120 negative samples, with three samples detected by NWLP only and one sample detected using our workflow only. Notably, these 4 samples showing a lack of concordance were all at the lowest limits of detection.

**Figure 5:**
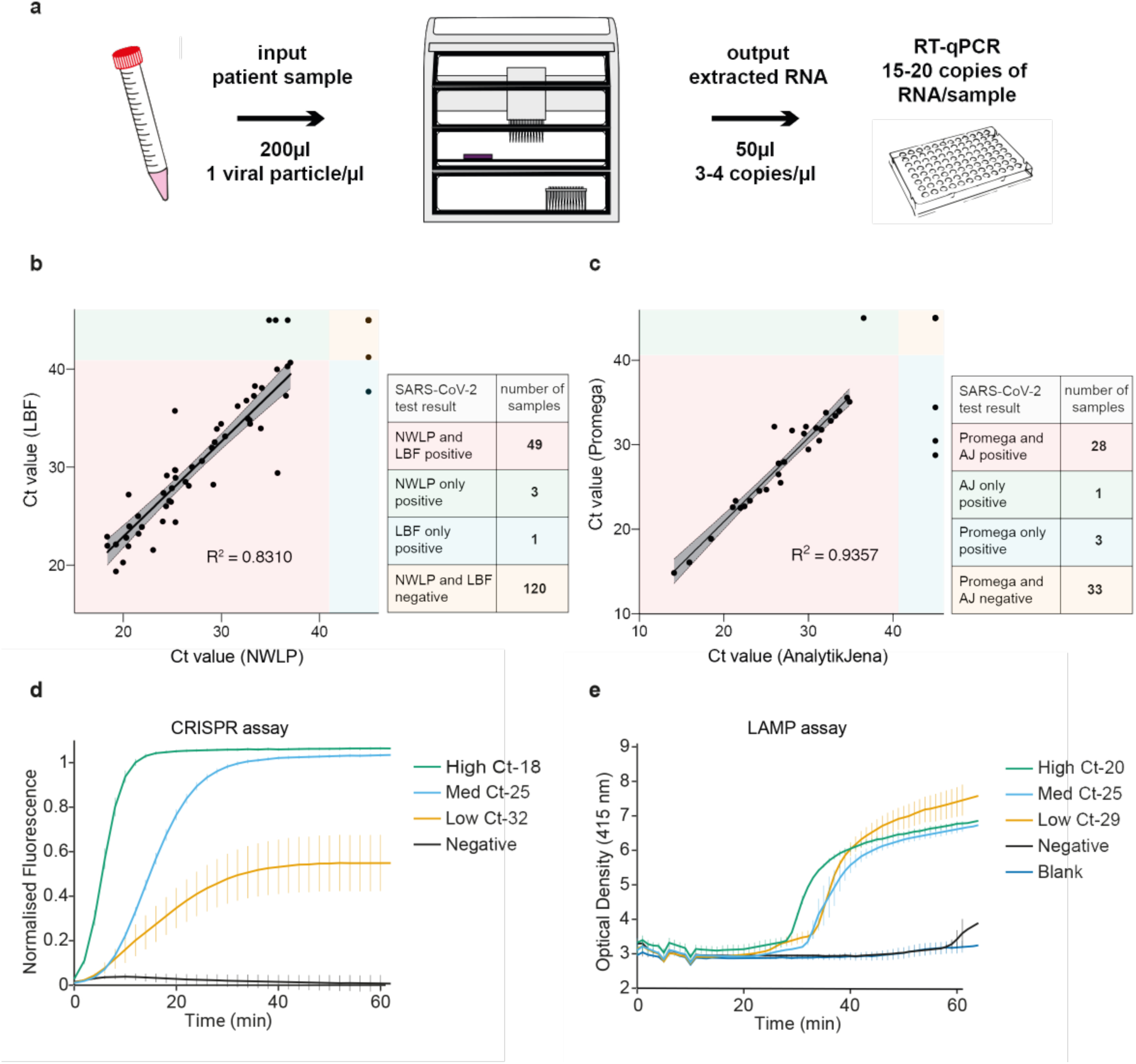
SARS-CoV-2 platform validation using patient samples. (**a**) Schematic of a typical workflow tracking viral copy number from a hypothetical patient sample where an input of 200 μL with a minimum 1 particle/ μL results in 3-4 copies of extracted RNA per microliter resulting in a range of 15-20 RNA copies per qPCR reaction. (**b**) Comparison of RT-qPCR Ct value results for the same 173 patient samples obtained by the North-West London Pathology (NWLP) laboratory and the London Biofoundry (LBF), based on the VLP workflow using the Analytik Jena RNA extraction kit and CDC N1 primer-probe set. (**c**) Validation of the Promega RNA extraction kit using 65 patient samples with the Analytik Jena RNA extraction workflow previously validated in (**b**). (**d**) Validation of the CRISPR NAT using patient samples at high, medium, and low SARS-CoV-2 concentrations as determined previously via RT-qPCR (Ct values of 18, 25, and 32 respectively). Error bars represent the standard error of the mean of three independent amplification replicates and four technical replicates for CRISPR detection. (**e**) Validation of the LAMP colorimetric NAT using patient samples at high, medium, and low SARS-CoV-2 concentrations as determined previously via RT-qPCR (Ct values of 20, 25, and 29 respectively). Error bars represent the standard error of the mean of three independent amplification replicates.

We then compared the Promega Maxwell HT Viral TNA extraction kit to the previously validated Analytik Jena innuPREP Virus DNA/RNA Kit workflow with a second set of patient samples. We observed high correlation (R^2^ = 0.9357) between Ct values for the same samples when processed with either the Promega or Analytik Jena extraction kits (Fig 5c). This highlights the strength of the platform providing consistent results for diverse reagent kits and supply chains. The more reagent kits are validated, the more resilience can be added through redundancy.

Finally, previously described detection assays [8], [17] were demonstrated in a HT-compatible format. Samples were extracted using the FeliX liquid handling protocol and resulting elution samples were transferred to plates certified for acoustic liquid handling. Miniaturised reactions were then set up to enable alternative detection modalities that are HT-compatible. The CRISPR-Cas workflow with RT-RPA amplification was shown to detect SARS-CoV-2 even at relatively low viral loads (Fig 5d). Additionally, the performance of colorimetric LAMP was successfully demonstrated using low, medium, and high viral load patient samples (Fig 5e).

## Discussion

In this study, we have been able to quickly repurpose automated liquid handling infrastructure in the London Biofoundry to establish two front-line SARS-CoV-2 testing platforms which are now operational in two London hospitals with a testing capacity of 2 000 samples per day. We have also developed CRISPR and colorimetric LAMP-based workflows and established a SARS-CoV-2 sVLP standard that had allowed us to validate the workflows within our biofoundry before implementation. During this process, we have identified a number of opportunities where biofoundries can be very effective in quickly providing increased SARS-CoV-2 testing capacity.

One major issue with standard diagnostic laboratory workflows is an over-reliance on a small number of manufacturers for infrastructure. For example, integrated platforms that allow HT sample processing – including automated patient sample nucleic acid extraction – are available from a few manufacturers, such as Roche, Abbott, Hamilton, Thermo Fisher, and Qiagen. However, at a time of unprecedented sample processing need, such as that imposed by the global COVID-19 pandemic, innovative approaches and non-traditional developers such as biofoundries, academic labs, start-ups, and SMEs can greatly expand testing options to add not only increased capacity, but also increased resiliency in mitigating supply chain bottlenecks [18], [19]. Biofoundries are agile facilities with a highly skilled workforce and cutting-edge equipment that can rapidly respond to such new challenges. Typically, they are not-for-profit institutions and therefore can evaluate different strategies unconstrained by commercial considerations. Their aim is to develop and apply purpose-built laboratory automation platforms, with an emphasis on versatile equipment, that can be adapted to a variety of synthetic biology workflows and support the translation of the latest scientific developments at their hosting research institutes and beyond. As lessons from management of the outbreak in Wuhan are beginning to emerge, it is becoming clear that automated diagnostic workflows, such as those implemented at the Huo-Yan diagnostic laboratory for processing 10 000 tests per day, play a pivotal role in containment of the virus [20].

When developing diagnostics for high-pressure pandemic scenarios, it is critical to create workflows that are modular and offer multiple contingency options, as reagent supply can quickly become a limiting factor to sample processing. Here we describe the rapid development of a HT diagnostic platform for the detection of SARS-CoV-2, using a synthetic VLP developed in-house under biosafety level 1 conditions. We use a versatile automated liquid handling device, the Analytik Jena CyBio FeliX, and validate 2 RNA extraction kits, multiple qPCR master mixes, as well as CRISPR- and colorimetric LAMP-based workflows, all in under four weeks. Importantly, we also validate the RNA extraction and RT-qPCR assay using patient samples, demonstrating a good correlation between a currently used clinical laboratory test for SARS-CoV-2 and our modular workflow. The framework provided for the validated platform may be further extended by alternative extraction and detection methodologies as well as in-house production and optimisation of kit components [21],[22]. This toolkit increases resilience of the SARS-CoV-2 NAT in case of shortages in extraction materials, RT-qPCR master mix, and laboratory equipment availability. The diagnostic output interchangeability is created not only by generating custom protocols for several commercially available kits, but also by adapting the CRISPR Cas13 detection and colorimetric LAMP systems to HT SARS-CoV-2 diagnostic testing. CRISPR-based detection technologies are also currently being developed by Sherlock Biosciences and Mammoth Biosciences to generate at-home point-of-care testing kits [14], [23], [24], as well as in CARMEN-Cas13, a microwell array which multiplexes virus detection [25].

Our workflow is easy to scale up, cost-effective and can provide similar output capacity to that offered by the ‘gold standard’ of commercial automated systems. For example, a single FeliX liquid handler and qPCR thermocycler can match the largest state-of-the-art Roche cobas^®^ 8800 platform, which can process 960 samples in eight hours. Additionally, excess viral RNA remaining from the Felix patient sample RNA extraction can be diverted to alternative analysis workflows such as next generation sequencing (NGS), which is not possible for some commercial platforms. Finally, our automated RNA extraction and qPCR workflow requires minimal specialist training and can be launched within one day. It is currently installed, and used in NHS diagnostic labs, where patient sample testing has been validated against large commercially available platforms, matching their precision and throughput.

Although NHS labs currently rely on qPCR workflows for all SARS-CoV-2 diagnostic testing, the potential of alternative detection technologies would allow for high-throughput testing for population screening and in low resource settings. Miniaturising LAMP and CRISPR reactions results in a slight loss of sensitivity and therefore may not be suitable for making diagnostic decisions where qPCR capacity is available, however, their isothermal incubation allows for thousands of samples to be tested simultaneously. LAMP is a particularly attractive technique because it has also been shown to be sensitive with heat inactivated samples, removing the bottleneck of RNA extraction [26]. Furthermore, these solutions can be deployed for testing in low-resource settings or at the point-of-care without expensive equipment requirements.

Synthetically engineered VLPs have been widely reported and commercially used as controls and standards in nucleic acid-based diagnostic tests (Asuragen), and have been developed as antigen epitopes in serological assays, where they are used to detect patient antibodies (Native Antigen) [27], [28]. MS2 VLPs carrying RNA payload, such as those used in this study for the detection of SARS-CoV-2 N gene RNA, provide a quick and reproducible system for generating extremely stable NAT controls. As such, we have purified and quantified large batches that are ready to be shared with, and employed by, others for diagnostic test development that relies on viral RNA detection. Furthermore, our VLP production and characterisation workflow can be modified to rapidly generate new controls mimicking emerging viral threats, thus enhancing preparedness for the development of new diagnostics in future epidemic or pandemic scenarios. In addition, automation equipment available in biofoundries can be used for large scale testing of antigen-presenting VLPs in developing antibody-based ELISA diagnostics as well as for performing high-throughput antiviral drug screens. The London Biofoundry is a founding member of the Global Biofoundry Alliance, which currently encompasses 26 such entities worldwide [29]. This network allows for easy sharing of reagents, protocols, and technical know-how, therefore automated diagnostic workflows developed by one partner can be quickly replicated around the world and increase capacity for testing and drug development to help counteract and prevent the global spread of emerging pathogens.

## Methods

### VLP preparation

The nucleic acid sequence of the N gene of SARS-CoV-2 (accession number: NC_045512) was ordered from GeneArt (ThermoFisher Scientific). The N gene was cloned into a previously described plasmid backbone (Addgene #128233) using Type IIs assembly. The sequence verified (Eurofins Genomics) plasmid was then transformed into Rosetta 2 (DE3) pLysS cells (Merck Millipore). An overnight culture was used to inoculated 200 mL of Terrific Broth (Merck) supplemented with 35 μg/mL of Chloramphenicol (Merck) and 50 μg/mL of Kanamycin (Merck) and grown at 37 °C, 200 rpm until an OD of 0.8. The culture was induced by supplementing with 0.5 mM IPTG (Merck) and grown at 30 °C for a further 16 hours. Cells were harvested at 4000 rpm at 4 °C and stored at −20 °C for later purification.

The protein purification workflow is based on previously described work [12], [30]. All protein purification steps were performed at 4 °C. The cell pellet was resuspended in 4 mL Sonication Buffer (50 mM Tris-HCl pH 8.0, 5 mM MgCl_2_, 5 mM CaCl_2_ and 100 mM NaCl) with 700 U RNase A (Qiagen), 2500 U BaseMuncher (Expedeon) and 200 U Turbo DNase (ThermoFisher Scientific). The cells were sonicated for a total of 2 minutes (50% amplitude, 30 seconds on, 30 seconds off) on wet ice. The lysate was then incubated for 3 hours at 37 °C. The lysate was centrifuged at 10 000 rpm for 10 minutes at room temperature in a microcentrifuge. The supernatant was then filtered with a 5 μm cellulose acetate (CA) filter before being mixed 1:1 with 2x Binding Buffer (100 mM Monosodium Phosphate Monohydrate pH 8.0, 30 mM Imidazole, 600 mM NaCl).

Supernatant was applied to a 5 mL HiTrap^®^ TALON^®^ crude column (GE Life Sciences) with a HiTrap^®^ Heparin HP column (GE Life Sciences) in series on an Äkta pure (GE Life Sciences) primed with Binding Buffer (50 mM Monosodium Phosphate Monohydrate pH 8.0, 15 mM Imidazole, 300 mM NaCl). The protein was eluted with a linear gradient of Elution Buffer (50 mM Monosodium Phosphate Monohydrate pH 8.0, 200 mM Imidazole, 300 mM NaCl) and then desalted and buffer exchanged into STE Buffer (10 mM Tris-HCl pH 7.5, 1 mM EDTA, 100 mM NaCl) using an Amicon Ultra-15 10K Centrifuge Filter (Merck). The protein concentration was measured using the Qubit Protein Assay Kit and Qubit 3 Fluorometer (ThermoFisher Scientific). The protein was then diluted in STE buffer, aliquotted and stored at -80°C.

### ddPCR

Droplet digital PCR was performed using the Bio-Rad QX200 Droplet Digital PCR system. Reactions were setup using the One-Step RT-ddPCR Advanced Kit for Probes (Bio-Rad) with primer and probe concentrations of 500 nM and 125 nM respectively. Data was exported in CSV format and analysed using a custom Python implementation (https://github.com/mcrone/plotlydefinerain) of an online tool (http://definetherain.org.uk). The online tool uses a positive control to define positive and negative droplets using K-means clustering, with rain being determined as anything outside three standard deviations from the mean of the positive and negative clusters. It then calculates final concentration based on the following equation.

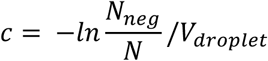

*c* = calculated concentration (copies/μL)

*N_neg_* = Number of negative droplets

*N* = Total number of droplets

*V_droplet_* = Average volume of each droplet (0.91 × 10^-3^ μL)

### DLS

Dynamic light scattering was performed using a Zetasizer Nano (Malvern Panalytical) according to the manufacturer’s instructions.

### qPCR

qPCR experiments were designed using the combination of SAS JMP and Riffyn. Primers, probes, and their relative concentrations were based on those recommended by the CDC and were ordered from IDT. TaqPath 1-Step RT-qPCR Master Mix (ThermoFisher Scientific), TaqMan Fast Virus 1-Step Master Mix (ThermoFisher Scientific), or Luna Universal Probe One-Step RT-qPCR (NEB) were used as the relevant master mix. qPCR reactions were otherwise set up according to the manufacturer’s instructions and thermocycling settings (annealing temperatures were according to the CDC protocol). Liquid transfers were performed using an Echo 525 (Labcyte). Plates were sealed with MicroAmp Optical Adhesive Films (ThermoFisher Scientific) and spun at 500 g in a centrifuge. An Analytik Jena qTower^3^ auto was used for thermocycling and measurements were taken in the FAM channel.

#### PCR Primers

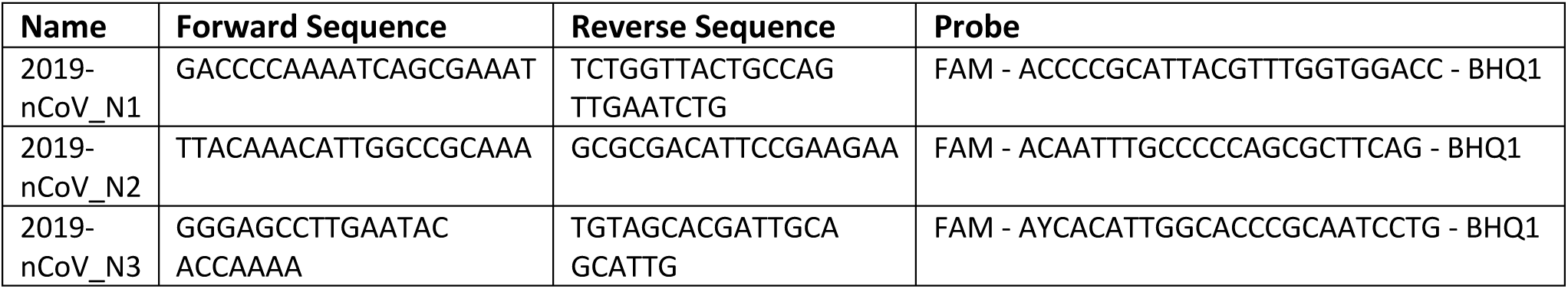

### LwCas13a Purification

LwCas13a was purified as described previously [14], with a few modifications. A plasmid expressing LwCas13 [pC013 - Twinstrep-SUMO-huLwCas13a was a gift from Feng Zhang (Addgene plasmid # 90097)] was transformed into Rosetta 2 (DE3) pLysS cells (Merck Millipore). An overnight culture was inoculated into 1 L of Terrific Broth (Merck) supplemented with 35 μg/mL of Chloramphenicol (Merck) and 50 μg/mL of Kanamycin (Merck) and grown at 37 °C, 160 rpm to an OD of 0.6. The culture was then induced with 0.5 mM IPTG (Merck), cooled to 18 °C and grown for a further 16 hours. Cells were harvested at 4000 rpm at 4 °C and stored at -20 °C for later purification.

All protein purification steps were performed at 4 °C. The cell pellet was resuspended in lysis buffer (20 mM Tris-HCl pH 8.0, 500 mM NaCl, 1 mM DTT) supplemented with protease inhibitors (cOmplete Ultra EDTA-free tablets, Merck) and BaseMuncher (Expedeon) and sonicated for a total of 90 seconds (amplitude 100% for 1 second on, 2 seconds off). Lysate was cleared by centrifugation for 45 minutes at 18 000 rpm at 4 °C and the supernatant was filtered through a 5 μm CA filter.

Supernatant was applied to a 5 mL StrepTrap HP column (GE Life Sciences) on an Äkta pure (GE Life Sciences). The buffer of the system was changed to SUMO digest buffer (30 mM Tris-HCL pH 8, 500 mM NaCl, 1 mM DTT, 0.15% Igepal CA-630). Sumo digest buffer (5 mL) supplemented with SUMO enzyme was then loaded directly onto the column and left to incubate overnight. The cleaved protein was then eluted with 5 mL of SUMO digest buffer. The elution fraction was diluted 1:1 with Ion Exchange low salt buffer (20 mM HEPES pH 7, 1 mM DTT, 5% Glycerol), applied to a Hitrap SP HP column (GE Life Sciences) and eluted using a gradient of the Ion Exchange high salt buffer (20 mM HEPES pH 7, 2000 mM NaCl, 1 mM DTT, 5% Glycerol). The eluted protein was then pooled, concentrated, and buffer exchanged into Storage buffer (50 mM Tris-HCl pH 7.5, 600 mM NaCl, 2 mM DTT, 5% Glycerol) using an Amicon Ultra-15 30K Centrifuge Filter (Merck). The protein concentration was measured using the Qubit Protein Assay Kit and Qubit 3 Fluorometer (ThermoFisher Scientific). The protein was then diluted, aliquoted and stored at -80 °C.

### gRNA Transcription and Quantification

DNA was ordered as ssDNA oligonucleotides from IDT and resuspended at 100 μM in Nuclease Free Duplex Buffer (IDT). Oligos contained a full-length reverse strand and a partial forward strand that contained only the T7 promoter sequence. Oligos were annealed by combining forward and reverse strands in equimolar concentrations of 50 μM and heating to 94 °C for 5 minutes and slow cooling (0.1°C/sec) to 25 °C in a thermocycler.

RNA was then *in vitro* transcribed using the TranscriptAid T7 High Yield Transcription Kit (ThermoFisher Scientific) according to the manufacturer’s instructions with a DNA template of 100 nM. Reactions were incubated for 16 hours at 37 °C. DNAse I was then added and incubated for 30 minutes at 37 °C.

Automated purification was performed using the CyBio FeliX liquid handling robot (Analytik Jena) using RNAClean XP beads (Beckman Coulter) according to the manufacturer’s instructions.

For automated quantification, samples were loaded into a 384 PP echo plate. Qubit Dye and Qubit Buffer were premixed at a ratio of 1:200 and loaded into a 6RES plate. Experimental design was performed using a custom Python script and Riffyn with each sample having four technical replicates that were randomly distributed in a Greiner 384 PS Plate (Greiner Bio-One). A standard curve of 9 concentrations (0, 5, 10, 15, 20, 40, 60, 80, 100 ng/μL) was obtained using the standards provided with the Qubit RNA BR Kit (ThermoFisher Scientific).

A volume of 9950 nL of the mix of Qubit Dye and Qubit buffer was added to each well using an Echo 525 (Beckman Coulter). A volume of 50 nL of sample was then added to each well using the Echo 525 (Beckman Coulter) and the plate was sealed with a Polystyrene Foil Heat Seal (4titude) using a PlateLoc Thermal Microplate Sealer (Agilent). Plates were centrifuged at 500 g for 1 minute before being kept in the dark for 3 minutes.

Plates were read using a CLARIOstar Plus (BMG Labtech) using the following settings: excitation wavelength of 625-15 nm, dichroic of 645 nm and emission of 665-15 nm and the Enhanced Dynamic Range (EDR) function. RNA molar concentration values were calculated, and the concentration was then normalised, RNA was aliquotted and subsequently stored at -80 °C.

#### gRNA Oligos

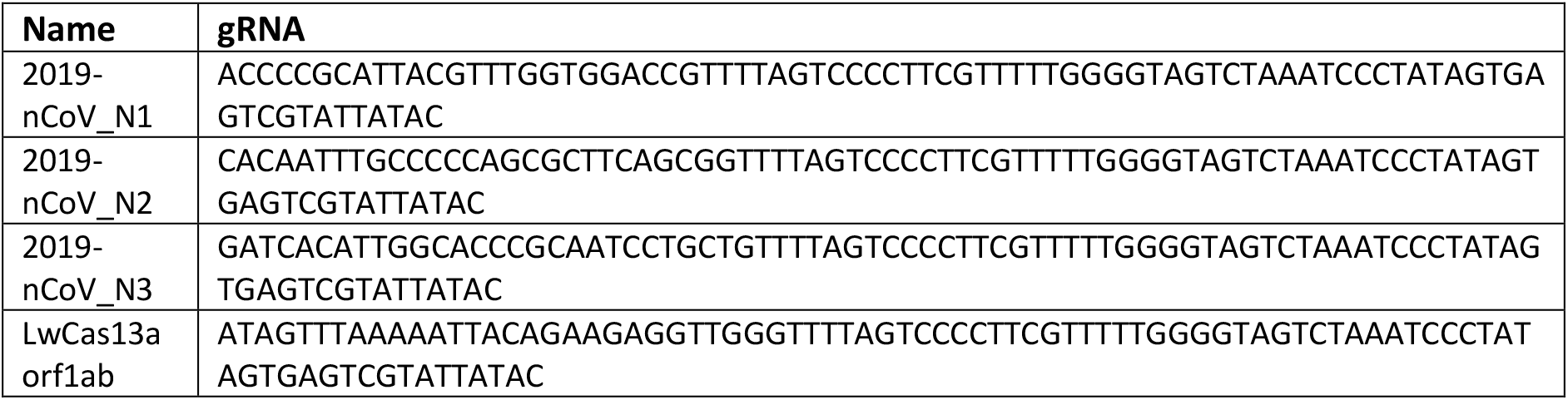

### CRISPR-Cas Assays with PCR amplification

Experiments were designed and randomised using JMP and Riffyn. Targets were pre-amplified using the Luna Universal One-Step RT-qPCR kit (NEB) with a primer concentration of 500 nM for 45 cycles.

All concentrations are final CRISPR reaction concentrations and the final CRISPR reaction volumes were 5 μL. An Echo 525 (Labcyte) was used to transfer CRISPR Master Mix (50 nM LwCas13a, 1 U/mL Murine RNAse inhibitor (NEB), 4 mM Ribonucleotide Solution Mix (NEB), 1.5 U/μl T7 RNA Polymerase (ThermoFisher Scientific) and 1.25 ng/μL HEK293F background RNA) in Nuclease Reaction Buffer (20 mM HEPES pH 6.8, 60 mM NaCl, 9 mM MgCl_2_) to a 384 well Small Volume LoBase Microplate (Greiner Bio-One). gRNA (25 nM) and a poly-U fluorescent probe (200 nM) were then added separately. An Echo 550 (Labcyte) was used to transfer pre-amplified products from a 384LDV Plus echo plate to initiate the reaction, the plate was sealed, spun at 500 g for 1 minute and read using a CLARIOstar Plus (BMG Labtech) with an excitation wavelength of 483-14 nm, emission of 530-30 nm, dichroic filter of 502.5 nm and enhanced dynamic range (EDR) enabled. Double orbital shaking of 600 rpm for 30 seconds was performed before the 1^st^ cycle. The reactions were incubated at 37 °C with readings taken every 2 minutes. Each reaction was normalised between a water input as 0 and an RNase I (ThermoFisher Scientific) input (0.25 U) as 1.

#### Amplification Primers

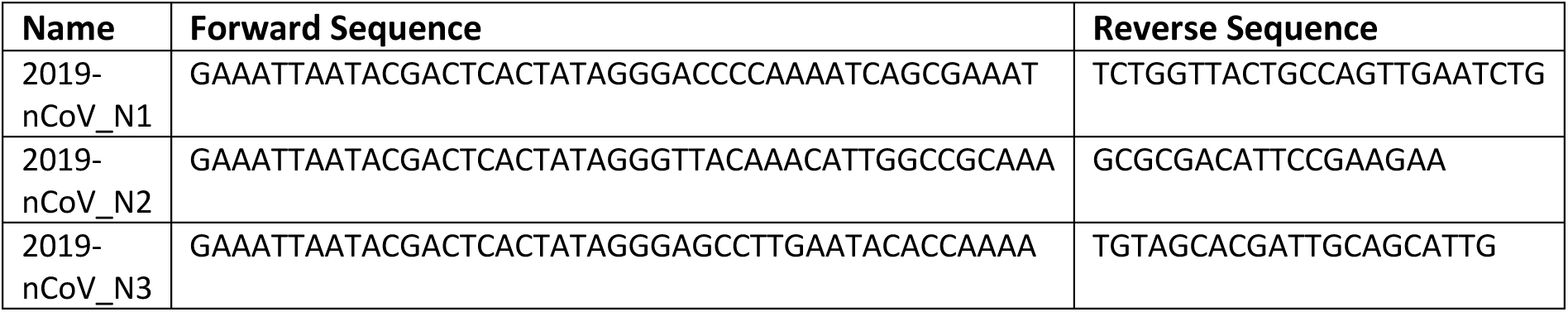

### Colorimetric LAMP Reactions with VLPs

Experiments were designed and randomised using JMP and Riffyn. Colorimetric LAMP reactions (NEB WarmStart^®^ Colorimetric LAMP 2X Master Mix) were performed as previously described [8] but with a lower final reaction volume of 5 μL. Master Mix, primers and template were transferred to a 384 Well Small Volume LoBase plate (Greiner Bio-One) using an Echo 525 (Labcyte). The plate was then sealed with a MicroAmp Optical Adhesive Film (ThermoFisher Scientific) and centrifuged for 1 minute at 500 g. The plate was incubated at 65 °C in a CLARIOstar Plus (BMG Labtech) and absorbance measurements were taken at 415 nm every minute for 60 minutes. Double orbital shaking of 600 rpm for 30 seconds was performed before the 1^st^, 6^th^, and 11^th^ cycles.

#### Primers

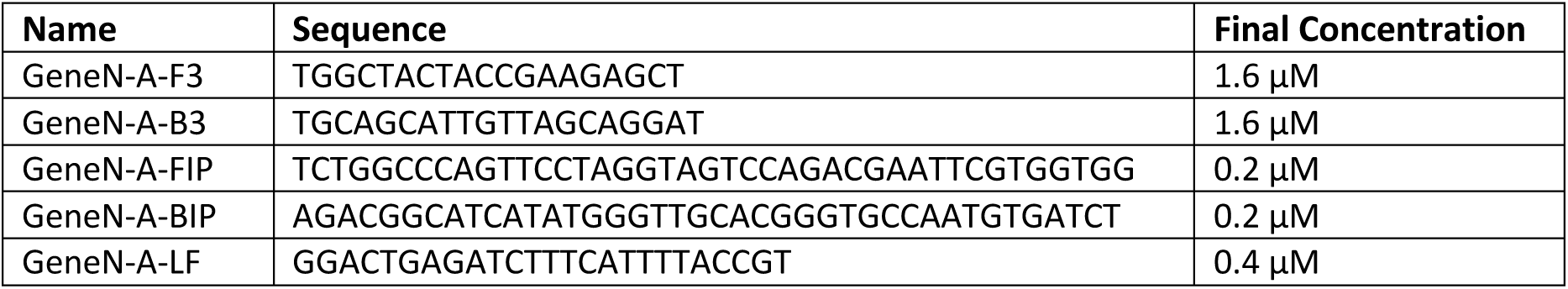
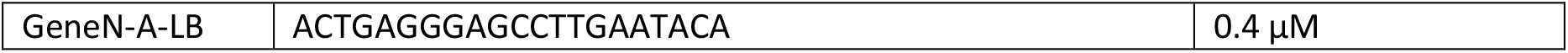

### RNA Extraction

RNA extraction was performed using a custom Analytik Jena CyBio FeliX script (available on request) for the Analytik Jena InnuPREP Virus DNA/RNA Kit – FX or the Promega Maxwell HT Viral TNA Kit. Samples of 200 μL were run and eluted in 50 μL of RNase Free Water.

### qPCR Patient Validation

Clinical material (Viral transport medium from throat/nose swabs), provided for validation by North West London Pathology (NWLP), included samples left over after clinical diagnosis as per standard practice for the validation of new assays and platforms. Results (Ct values) were compared directly with those obtained by NWLP.

qPCR reactions were setup using the TaqPath 1-Step RT-qPCR Master Mix, CG kit, and the CDC N1 Primers according to the manufacturer’s instructions and thermocycling settings (annealing temperatures were according to the CDC protocol). Final reaction volumes were 10 μL with 5 μL of extracted RNA template. Liquid transfer of the qPCR master mix was performed using an Echo 525 (Labcyte) from a 6 well reservoir (Labcyte). Extracted RNA templates were transferred using a multichannel pipette. Plates were sealed with MicroAmp Optical Adhesive Films (ThermoFisher Scientific) and spun at 500 g in a centrifuge. An Analytik Jena qTower^3^ auto was used for thermocyling and measurements were taken in the FAM channel.

### CRISPR-Cas Assays with RT-RPA amplification

Experiments were designed and randomised using JMP and Riffyn. Targets were pre-amplified using the TwistAmp Liquid Basic Kit (TwistDx) supplemented with 0.5 U/μL Murine RNase Inhibitor (NEB) and 0.08 U/μL Omniscript (Qiagen). Final reactions had a final volume of 14 μL and were setup in Echo 384 LDV Plus plates (final primer concentration of 0.45 μM and 2 μL of purified patient RNA template). All concentrations are final CRISPR reaction concentrations and the final CRISPR reaction volumes were 5 μL. An Echo 525 (Labcyte) was used to transfer CRISPR Master Mix (50 μM LwCas13a, 1 U/μL Murine RNase inhibitor (NEB), 4 mM Ribonucleotide Solution Mix (NEB), 1.5 U/μl T7 RNA Polymerase (ThermoFisher Scientific) and 1.25 ng/μL HEK293F background RNA) in Nuclease Reaction Buffer (20 mM HEPES pH 6.8, 60 mM NaCl, 9 mM MgCl_2_) to a 384 well Small Volume LoBase Microplate (Greiner Bio-One). gRNA (25 nM) and a poly U fluorescent probe (200 nM) were then added separately. An Echo 550 (Labcyte) was used to transfer pre-amplified products (250 nL) from the 384LDV plus echo plate to initiate the reaction, the plate was sealed, centrifuged at 500 g for 1 minute and read using a CLARIOstar Plus (BMG Labtech) with an excitation wavelength of 483-14 nm, emission of 530-30 nm,dichroic filter of 502.5 nm and enhanced dynamic range (EDR) enabled. Double orbital shaking of 600 rpm for 30 seconds was performed before the 1^st^ cycle. The reactions were incubated at 37 °C with readings taken every 2 minutes. Each reaction was normalised between a water input as 0 and an RNase I (ThermoFisher Scientific) input (0.25 U) as 1. Error was calculated as the standard error of three amplification and four CRISPR replicates.

#### Amplification Primers

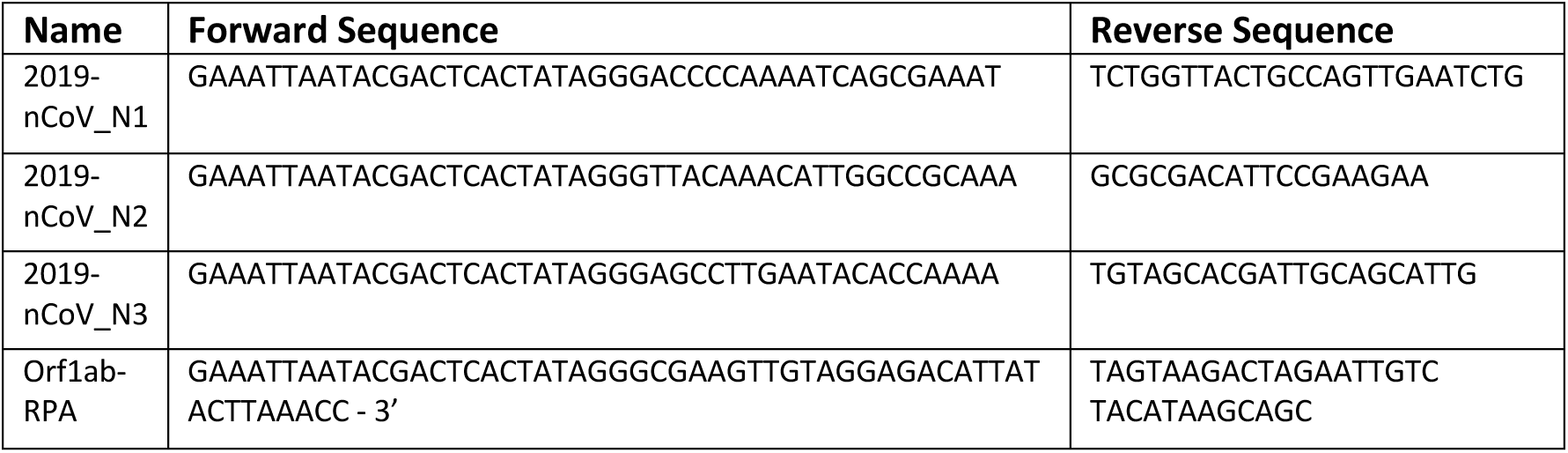

### Colorimetric LAMP Reactions with patient samples

Experiments were designed and randomised using JMP and Riffyn. Colorimetric LAMP reactions (NEB WarmStart^®^ Colorimetric LAMP 2X Master Mix) were performed as previously described [8] but with a lower final reaction volume of 5 μL and template of 2 μL. Master Mix, primers and template were transferred to a 384-Well Small Volume LoBase plate (Greiner Bio-One) using an Echo 525 and Echo 550 (Labcyte). The plate was then sealed with a MicroAmp Optical Adhesive Film (ThermoFisher Scientific) and centrifuged for 1 minute at 500 g. The plate was incubated at 65 °C in a CLARIOstar Plus (BMG Labtech) and absorbance measurements were taken at 415 nm every minute for 60 minutes. Double orbital shaking of 600 rpm for 30 seconds was performed before the 1^st^, 6^th^, and 11^th^ cycles.

#### Primers

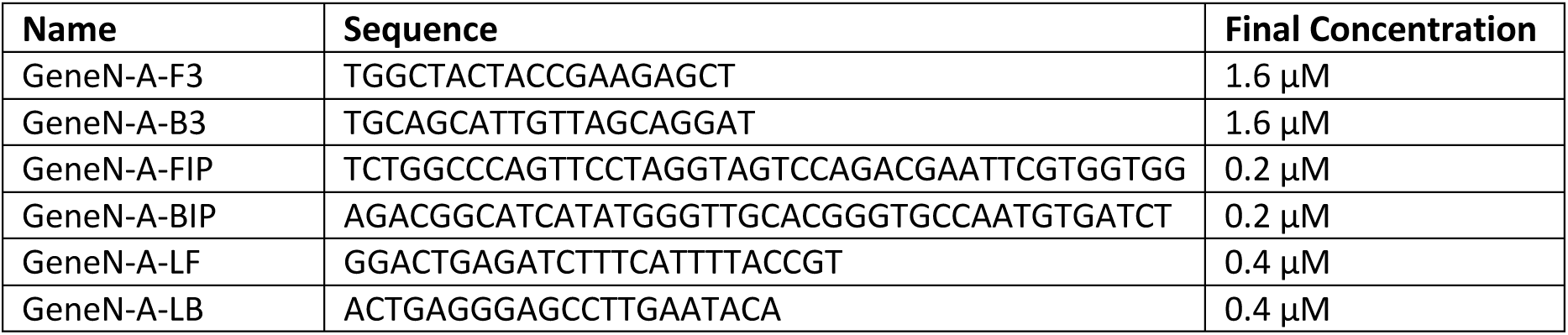

### Ethics statement

Surplus clinical material was used to validate the assay as per normal practice and does not require ethical review.

## Data Availability

All data is available upon request.

## Acknowledgements

We would particularly like to thank the UK Dementia Research Institute for rapidly funding the initial workflow development and for continuing support via the DRI Care Research and Technology Centre at Imperial College. We also acknowledge funding from UKRI-EPSRC (EP/R014000/1, EP/S001859/1), UKRI-BBSRC (BB/M025632/1) and the National Physical Laboratory (NPL). We would like to thank Graham Taylor, Myra McClure, Arthi Anand, and Panagiotis Pantelidis for their excellent clinical diagnostic guidance during this project and support with the surplus clinical material. We also thank Andrew Griffiths at the DRI Care Research and Technology Centre for project management support. We would also like to thank Analytik Jena and in particular Debra Conway and BMG Labtech for providing equipment and support throughout the process. We also thank Professor Charles Bangham for the use of the QX200 ddPCR setup. MiC would like to thank the Science Team at Riffyn for training and continued support. We thank Matthew Haines in the Freemont lab for critical reading of the manuscript.

